# Electronic cigarette communications between patients and physicians in the United States

**DOI:** 10.1101/2021.11.01.21265609

**Authors:** Cristine D. Delnevo, Michelle Jeong, Arjun Teotia, Michelle M. Bover Manderski, Binu Singh, Mary Hrywna, Michael B. Steinberg

## Abstract

**Importance:** Physicians play a primary role in smoking cessation, and their communication regarding e-cigarettes needs to be understood.

**Objective:** To examine physician-patient communication regarding e-cigarettes.

**Design:** A national, repeated cross-sectional survey in 2018 and 2019 was conducted.

**Setting:** Participants were invited by mail; surveys were completed online.

**Participants:** Response rates were 51.8% (2018) and 59.1% (2019), resulting in 2,058 board-certified physicians from family medicine, internal medicine, obstetrics/gynecology, cardiology, pulmonary/critical care, and hematology/oncology.

**Exposures:** Physician demographics, tobacco use, medical specialty, and harm-reduction beliefs (i.e., not all tobacco products equally harmful); two hypothetical clinical scenarios.

**Main outcomes and measures:** Physicians’ self-reported e-cigarette communication behaviors (being asked about e-cigarettes by patients and recommending e-cigarettes to patients), and hypothetical e-cigarette communication in two clinical scenarios.

**Results:** Among 2,058 physicians, the mean age was 51.6 years and 41.5% were female. Over 60% of physicians believed all tobacco products are equally harmful. Overall, 69.8% of physicians reported ever being asked about e-cigarettes by their patients (35.9% in the past 30 days), while 21.7% reported ever recommending e-cigarettes to a patient (9.8% in the past 30 days). Pulmonologists (adjusted odds ratio [AOR], 2.14, 95% CI, 1.10-4.16) and cardiologists (AOR, 2.04; 95% CI, 1.03-4.05), as well as physicians who implemented the US Public Health Service Clinical Practice Guidelines (AOR, 1.77; 95% CI, 1.12-2.80) had greater odds of recommending e-cigarettes to patients. Physicians who endorsed a harm-reduction perspective (AOR, 3.04, 95% CI, 2.15-4.31) and had ever smoked cigarettes (AOR, 1.98; 95% CI, 1.27-3.08) were significantly more likely to recommend e-cigarettes. Being asked about e-cigarettes by patients was a strong predictor of physicians’ recommending (AOR,16.6; 95% CI, 10.3-26.7). In clinical scenarios, physicians were overall more likely to recommend e-cigarettes for cessation to an older, heavy smoker with multiple unsuccessful quit attempts than a younger, light smoker with no prior cessation treatments (49.3% vs. 15.2%, *p*<.001).

**Conclusions and relevance:** Findings suggest physicians may recommend e-cigarettes for cessation under certain circumstances. Given the role of e-cigarettes in FDA’s comprehensive nicotine policy, there is need for continued physician education regarding e-cigarette efficacy, particularly correcting misperceptions regarding harm reduction.

**Key Points:** *Question:* How do physicians communicate with their patients about e-cigarettes?

*Findings:* In this repeated cross-sectional survey of 2,058 respondents, physicians who were asked about e-cigarettes by their patients, endorsed a harm-reduction perspective, or had ever smoked were significantly more likely to recommend e-cigarettes to patients. Clinical scenarios showed physicians were significantly more likely to recommend e-cigarettes for an older, heavy smoker with prior unsuccessful quit attempts, and use of FDA-approved pharmacotherapy for a younger, light smoker with no prior cessation treatments.

*Meaning:* Physicians may recommend e-cigarettes for cessation under certain circumstances, warranting continued physician education regarding e-cigarette efficacy.

## Introduction

Although tremendous progress has been made in reducing smoking over the last several decades, smoking continues to cause approximately 480,000 deaths in the United States each year,^1^ and effective strategies to reduce this loss are still needed. The 2018 report by the National Academies of Sciences, Engineering, and Medicine demonstrated varying levels of evidence regarding the risks to youth and the potential benefits of e-cigarettes, such as smoking cessation and harm reduction for adult cigarette smokers.^2^ While a high number of youth use e-cigarettes,^3^ the data suggest that the majority of adult e-cigarette users are current and former cigarette smokers who turned to e-cigarettes as a less risky alternative to smoking and for smoking cessation, despite limited evidence on safety and efficacy.^4-9^ The most recent systematic review on the use of e-cigarettes for smoking cessation concluded that use of nicotine-containing e-cigarettes was associated with significantly higher quit rates when compared to NRT or behavioral support.^10^ While the long-term safety of these devices remains unknown, it is likely that they are far less harmful than combusted cigarettes.^11^ For these reasons, e-cigarettes remain a potentially beneficial option for current adult cigarette smokers who are unable or unwilling to quit otherwise.

Physicians play an important role in smoking cessation as they treat adult smokers on a regular basis and have established provider-patient relationships.^12^ Many physicians report being asked by their patients about e-cigarettes,^13-15^ which is not surprising given that patients who smoke cite physicians as a trustworthy source of e-cigarette information.^16^ However, how physicians respond to such patient questions is less clear. A recent review of the research literature on health care providers and e-cigarettes found that providers, including physicians, held mixed views regarding the safety and role of e-cigarettes in smoking cessation, which was partially explained by different e-cigarette messaging across countries.^17^ Indeed, in stark contrast to the United States,^18,19^ physicians in the United Kingdom are encouraged to consider e-cigarettes as an option for smoking cessation and to support smokers who wish to try them.^20-22^

Additionally, the few physician studies in the US regarding e-cigarette communications suggest that while physicians were unlikely to recommend e-cigarettes for smoking cessation,^15,23^ they are open to recommending e-cigarettes in the future if data becomes available suggesting effectiveness.^24^ Although some early data exist regarding e-cigarette physician-patient communications in the US, with one exception^14^ the US studies were limited to local convenience samples^25-27^ and/or single specialties.^28-30^ Furthermore, these products, the scientific evidence base, and the policy environment surrounding e-cigarettes are evolving. The aim of this study is to add to the growing body of knowledge about physician-patient communications and recommendations of e-cigarettes using two waves of a large national survey of physicians in the US.

## Methodology

This study was a national, repeated cross-sectional, web-push mail survey of physicians in 2018 and 2019. The sampling frame was compiled from the American Medical Association’s master list, purchased from Medical Marketing Services (MMS), and included a random sample of board-certified physicians practicing in various specialties. In each year, a total of 3,000 physicians were sampled. In 2018, the sample was distributed evenly across six specialties of interest (i.e., 500 were randomly selected from each specialty): family medicine, internal medicine, obstetrics and gynecology, cardiology, pulmonology, and oncology. In 2019, the sample was distributed evenly across four specialties of interest (i.e., 750 randomly selected from each specialty): family medicine, internal medicine, obstetrics and gynecology, and pediatrics. Pediatricians were added in 2019 given the rise in e-cigarette use among youth.

Survey fielding occurred from February to July 2018 (Wave 1) and April to July 2019 (Wave 2). Survey method experiments were embedded in the study in Wave 1 to refine the web-push field procedures^31^ and test the impact of differing survey incentives ($25 vs $50); Wave 2 used a $50 incentive. An initial mailing contained a personalized introductory cover letter, an upfront incentive, and instructions on how to complete the web survey (i.e., the survey URL was provided with an anonymous login code). After 1 week from the first mailing, a second mailing contact by postcard was sent to nonrespondents. The third mailing to nonrespondents mirrored the first contact, excluding the gift card. The fourth mailing to all nonrespondents included a paper survey as well as a cover letter with instructions on how to complete the web survey, allowing nonrespondents on the fourth contact to choose data collection mode (i.e., paper survey or web survey).

Using AAPOR’s response rate 3 calculation,^32^ which estimates the proportion of cases of unknown eligibility that are eligible, our response rate was 51.8% in 2018 and 59.1% in 2019. Given the focus of this manuscript on patient–provider communications about electronic cigarettes in the context of adult cigarette smoking, the analytic sample excluded pediatricians who answered different questions regarding e-cigarettes (i.e., prevention focused).

The survey instrument was developed using key domains of interest including but not limited to demographics, medical background, clinical practice, tobacco treatment practices, harm reduction beliefs, e-cigarette patient communication and messaging. Survey items were adapted from previous physician surveys.^14,33^ The results of our previous qualitative study of physicians also informed question development^24^ and the instrument underwent cognitive testing. Physicians were presented with two clinical scenarios and then asked how they would communicate with that specific patient about e-cigarettes. The first patient was a young female, light smoker who had never tried any cessation methods while the other patient was an older male, heavy smoker with multiple unsuccessful quit attempts using various cessation medications.

Logistic regression analyses were used to examine factors associated with physicians being asked by patients about e-cigarettes (yes v. no) and physicians recommending electronic cigarettes (yes v. no). Paired t-tests were used to compare physician responses to two clinical scenarios. Analyses were performed using Stata MP17.

The Rutgers Biomedical Health Sciences Institutional Review Board approved the study as exempt. This study followed the American Association for Public Opinion Research (AAPOR) reporting guideline.

## Results

The sample contains cross-sectional data from 2058 respondents over two waves (2018 and 2019). Table 1 summarizes the demographic and tobacco-related characteristics of our respondents. Overall, 58.5% of the respondents were male, with a mean age of 51.6 years (min = 30, max = 85) and 66% of the respondents were Non-Hispanic Whites, 12.8% were Asian/Pacific Islanders, 6.9% were South Asians. With respect to medical specialty, since both waves included primary care physicians, they constituted the majority of the sample, with 28.1% specializing in family medicine, followed by obstetrics/gynecology (27%), and internal medicine (22.3%), whereas sub-specialists, who only participated in Wave 1, made up a smaller percent as follows: pulmonary (8.6%), cardiology (7.2%), and oncology (6.9%).

**Table 1.** Demographics.

|  | Distribution |  |
| --- | --- | --- |
|  | N | % |
| Overall | 2058 | 100.0 |
| Age, mean ± SD | 1943 | 51.6 (10.5) |
| Gender |  |  |
| Male | 1173 | 58.5 |
| Female | 831 | 41.5 |
| Race/Ethnicity |  |  |
| NH White | 1318 | 66.1 |
| NH Black | 96 | 4.8 |
| Hispanic | 88 | 4.4 |
| Asian/Pacific Islander | 256 | 12.8 |
| South Asian | 138 | 6.9 |
| NH Other | 99 | 5.0 |
| Specialty |  |  |
| Family Medicine | 574 | 28.1 |
| Ob/Gyn | 551 | 27.0 |
| Internal Medicine | 455 | 22.3 |
| Pulmonary | 175 | 8.6 |
| Cardiology | 147 | 7.2 |
| Oncology | 140 | 6.9 |
| Year of Data Collection |  |  |
| Wave 1 /2018 | 1,058 | 51.4 |
| Wave 2 /2019 | 1,000 | 48.6 |
| PHS Awareness |  |  |
| Unaware | 650 | 32.5 |
| Aware, not read | 811 | 40.6 |
| Read, not used | 146 | 7.3 |
| Uses | 392 | 19.6 |
| Risk Continuum |  |  |
| All forms of tobacco equally harmful | 1,222 | 60.1 |
| Cigarettes are the most dangerous | 810 | 39.9 |
| Ever Smoker |  |  |
| Yes | 253 | 12.5 |
| No | 1,765 | 87.5 |
| Ever E-Cig User |  |  |
| Yes | 71 | 3.5 |
| No | 1,946 | 96.5 |

Among all respondents, one out of three reported they were unaware of Public Health Service Clinical Practice (PHS) guidelines on tobacco at the time of the survey, while 19.6% reported using the guidelines. When asked about harm reduction beliefs, the majority of physicians (60.1%) endorsed the belief that all forms of tobacco were equally harmful and cessation from all tobacco was the best approach whereas 39.9% endorsed the belief that getting smokers to stop smoking cigarettes should be the goal, even if it meant switching to less harmful forms of tobacco. Lifetime cigarette smoking among physicians was lower than the national average, with 12.5% reporting having smoked 100 cigarettes in their lifetime and 3.5% indicated they had tried an e-cigarette.

Table 2 details physician-patient communications regarding e-cigarettes. Overall, 69.8% of physicians reported ever being asked about e-cigarettes by their patients and 35.9% reported being asked about them in the 30 days preceding the survey. Additionally, 21.7% of physicians reported ever recommending e-cigarettes to a cigarette smoker, and 9.8% reported making such a recommendation in the past 30 days. Overall, males were more likely to report being asked about e-cigarettes in the past 30 days (40.3% v. 30.5% for females). Males were also more likely to report having recommended e-cigarettes in the past 30 days (12.1% v. 6.6% for females). Black physicians were least likely to report ever being asked about e-cigarettes by their patients (49.5%) whereas Hispanic physicians were most likely to report being asked about e-cigarettes within the past 30 days (47.7%). There were no significant differences in recommendations by physician race/ethnicity. Pulmonologists and family medicine physicians were more likely to report having ever been asked about e-cigarettes than other specialists (90.3% and 84.6%) as well as in the past 30 days (59.4% and 47.6%). Pulmonologists were also most likely to recommend e-cigarettes in the past 30 days, followed by cardiologists and family medicine (17.2%, 12.9%, and 10.6%). Interestingly, while cardiologists were less likely to be asked about e-cigarettes than family medicine, our data showed that cardiologists were more likely to recommend e-cigarettes to patients in the past 30 days.

**Table 2.** Patient prompting about e-cigarettes and physician e-cigarette recommendation (2018-2019)

|  | Ever asked about e-cig by patient |  | Asked about e-cig by patient in P30D |  | Ever recommended ecig to a patient |  | Recommended ecig to a patient in P30D |  |
| --- | --- | --- | --- | --- | --- | --- | --- | --- |
|  |  | p-value |  | p-value |  | p-value |  | p-value |
| Overall | 69.8% |  | 35.9% |  | 21.7% |  | 9.8% |  |
| Age, mean ± SD | | | | | | | | |
| Gender |  |  |  |  |  |  |  |  |
| Male | 73.0% | 0.00 | 40.3% | <.0001 | 25.2% | <.0001 | 12.1% | <.0001 |
| Female | 65.9% |  | 30.5% |  | 17.2% |  | 6.6% |  |
| Race/Ethnicity |  |  |  |  |  |  |  |  |
| NH White | 73.1% | <.0001 | 38.6% | <.0001 | 23.6% | *ns* | 10.5% | *Ns* |
| NH Black | 49.5% |  | 24.0% |  | 16.7% |  | 6.3% |  |
| Hispanic | 73.9% |  | 47.7% |  | 15.9% |  | 6.8% |  |
| Asian/Pacific Islander | 60.9% |  | 23.6% |  | 17.9% |  | 7.8% |  |
| South Asian | 72.5% |  | 39.9% |  | 21.2% |  | 11.0% |  |
| NH Other | 63.6% |  | 32.3% |  | 21.2% |  | 11.1% |  |
| Specialty |  |  |  |  |  |  |  |  |
| Ob/Gyn | 50.1% | <.0001 | 20.9% | <.0001 | 15.1% | <.0001 | 7.1% | 0.001 |
| Cardiology | 72.1% |  | 30.3% |  | 24.8% |  | 12.9% |  |
| Family Medicine | 84.6% |  | 47.6% |  | 24.9% |  | 10.6% |  |
| Internal Medicine | 72.5% |  | 36.8% |  | 22.7% |  | 9.5% |  |
| Oncology | 52.9% |  | 22.3% |  | 14.4% |  | 5.0% |  |
| Pulmonary | 90.3% |  | 59.4% |  | 33.1% |  | 17.2% |  |
| PHS Awareness |  |  |  |  |  |  |  |  |
| Unaware | 64.6% | <.0001 | 29.8% | <.0001 | 18.3% | 0.01 | 7.7% | 0.023 |
| Aware, not read | 67.8% |  | 32.8% |  | 22.5% |  | 9.5% |  |
| Read, not used | 71.2% |  | 42.5% |  | 20.1% |  | 10.4% |  |
| Uses | 83.4% |  | 52.6% |  | 27.0% |  | 13.6% |  |
| Risk Continuum |  |  |  |  |  |  |  |  |
| All forms of tobacco equally harmful | 67.9% | 0.01 | 35.1% | *ns* | 13.3% | *p<.0001* | 6.0% | <.0001 |
| Cigarettes are the most dangerous | 73.1% |  | 37.2% |  | 34.1% |  | 15.1% |  |
| Ever Smoker |  |  |  |  |  |  |  |  |
| Yes | 70.4% | *ns* | 36.5% | *ns* | 28.8% | 0.01 | 16.7% | <.0001 |
| No | 69.7% |  | 36.1% |  | 20.8% |  | 8.6% |  |
| Ever E-Cig User |  |  |  |  |  |  |  |  |
| Yes | 73.2% | *ns* | 43.7% | *ns* | 40.0% | *p<.0001* | 18.6% | 0.01 |
| No | 69.6% |  | 35.8% |  | 21.1% |  | 9.3% |  |
| Year of Data Collection |  |  |  |  |  |  |  |  |
| Wave 1 /2018 | 69.9% | *ns* | 35.2% | *ns* | 22.2% | *ns* | 10.0% | *Ns* |
| Wave 2 /2019 | 69.7% |  | 36.6% |  | 21.2% |  | 9.5% |  |

Physicians unaware of PHS guidelines were less likely to recommend e-cigarettes to patients, with no significant difference in being asked about e-cigarettes in the past 30 days across PHS categories. While there were no meaningful relationships between harm reduction beliefs and being asked about e-cigarettes by patients, the relationship between physicians’ harm reduction beliefs and their e-cigarette recommendation practices was significant. Physicians who believed that cigarettes were the most dangerous tobacco product were significantly more likely to report ever recommending e-cigarettes (34.1% v. 13.3%) as well as in the past 30 days (15.1% v. 6%), compared to physicians who believed all forms of tobacco were equally harmful. Physicians who reported ever smoking, as well as trying e-cigarettes, were more likely to recommend e-cigarettes compared to non-smokers or those that had not tried e-cigarettes.

Predictors of being asked about e-cigarettes by patients are found in Table 3. Increasing physician age and Asian/Pacific Islander descent was significantly associated with being less likely to report being asked by their patients about e-cigarettes, and male physicians were significantly more likely to report being asked about e-cigarettes. Specialty was significantly associated with reporting being asked about e-cigarettes: pulmonologists had the highest odds of being asked, followed by family medicine, internal medicine, and cardiologists. Physician awareness and implementation of the PHS guidelines was a robust and significant predictor of physicians reporting they were asked about e-cigarettes. Lastly, being asked about e-cigarettes in the 30 days preceding the survey was significantly higher in 2019 than in 2018.

**Table 3.**
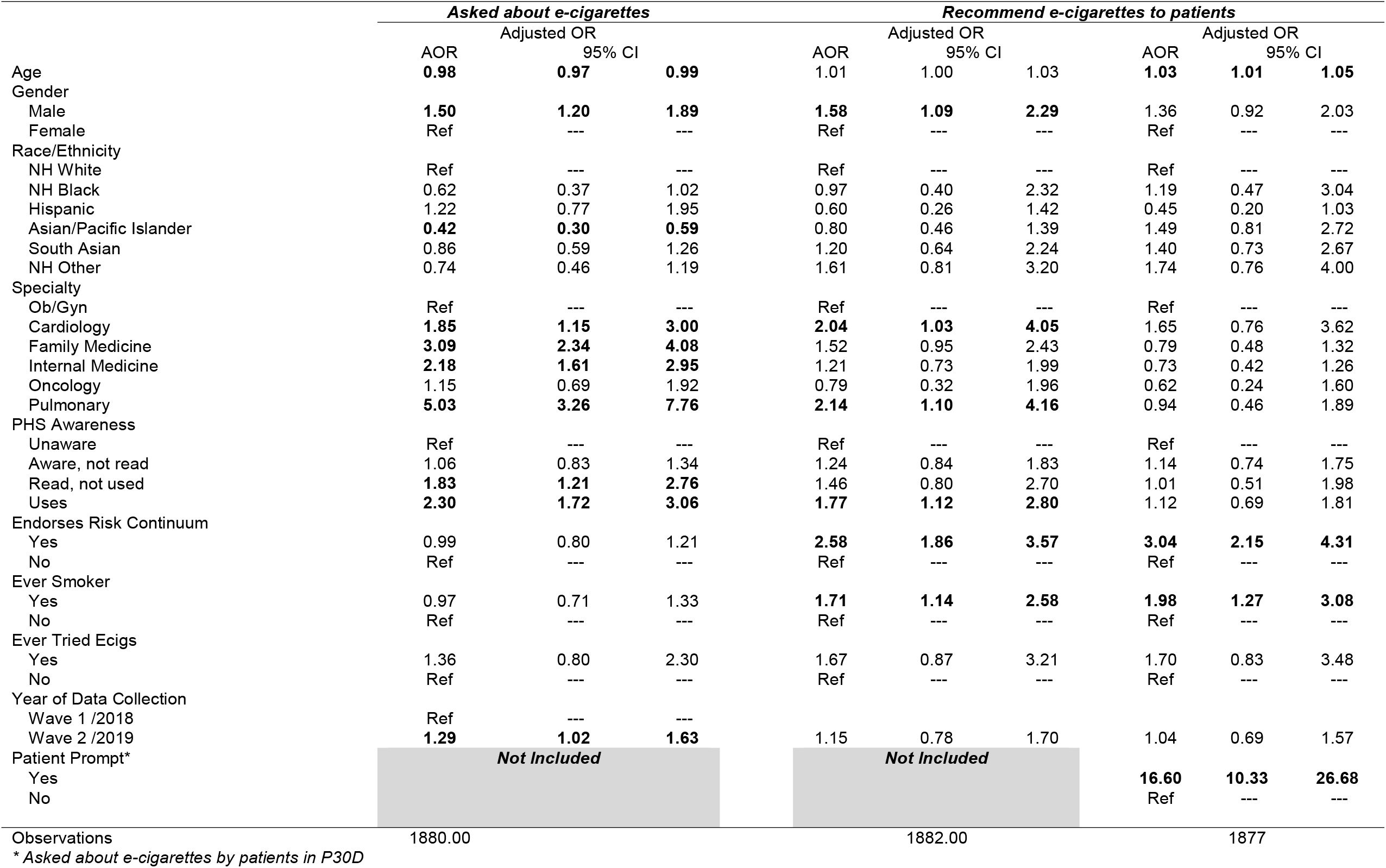
predictors of being asked by patients about e-cigarettes and recommending e-cigarettes to patients in past 30 days.

With respect to recommending e-cigarettes to their patients, we ran two models (see Table 3), one of which controlled for whether or not a patient had asked the physician about e-cigarettes or “patient prompting.” When patient prompting (AOR=16.6, 95% CI 0.3-26.7) was added to the model, male sex, physician specialty and engagement with the PHS guidelines were no longer significant predictors of e-cigarette recommendations. Having ever been a regular smoker and endorsing a harm-reduction perspective remained significant predictors of e-cigarette recommendations, even when controlling for patient prompting.

Table 4 highlights differences in physician communication on the patient clinical scenario by medical specialty. Overall, physicians were most likely to communicate that they preferred the patient to use FDA-approved pharmacotherapy over an e-cigarette to help them quit. This message wass endorsed more often when the patient was a young female, light smoker (82.1%) than an older male, heavy smoker with multiple failed quit attempts (56.8%). Likewise, physicians were more likely to encourage smokers to try e-cigarettes to transition off cigarettes when the patient was a heavy smoker with multiple failed quit attempts (49.3%) than if the patient had never tried quitting before (15.2%). Relative to other specialists, cardiologists were most likely to recommend e-cigarettes to the younger patient (23.1%). Roughly one in four physicians communicated that e-cigarettes were harmful and discouraged use; this was significantly more likely with the younger light smoker than the older heavier smoker (31.7% vs. 24.9%). This tailored, risk-based communication was consistent across medical specialties. Lastly, there were no significant differences in physician communication that e-cigarettes were not effective for cessation by patient type.

**Table 4.**
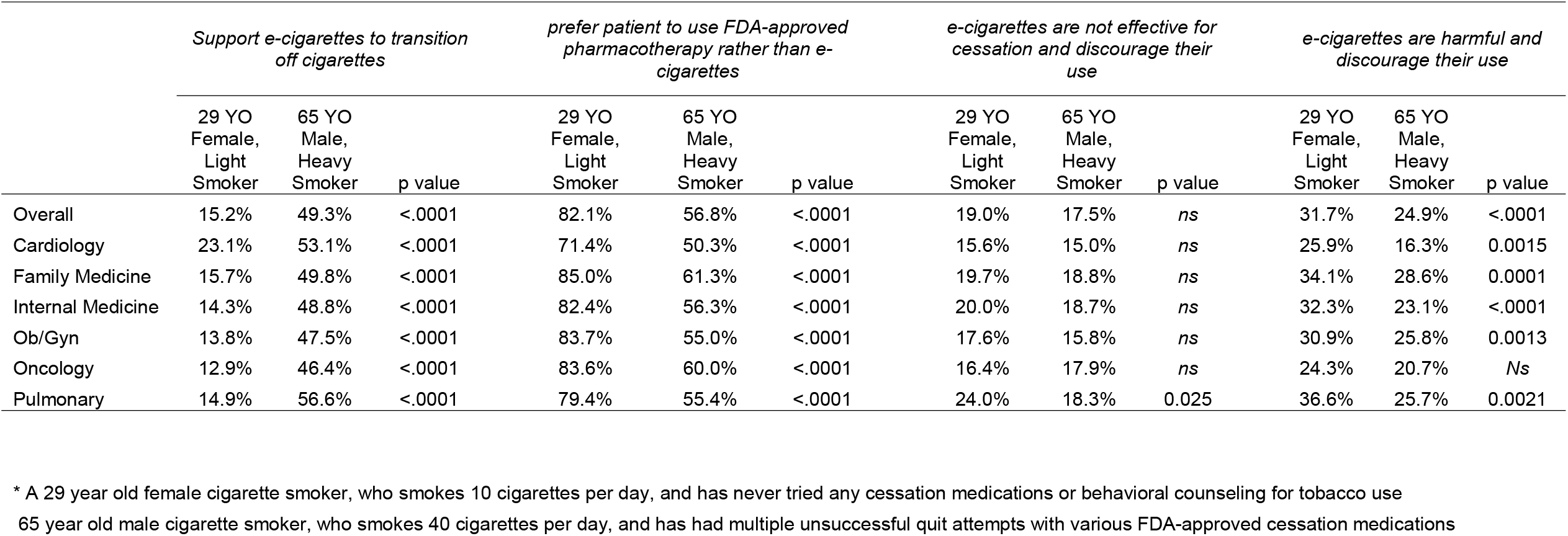
Messaging to smokers regarding e-cigarettes by medical specialty and patient characteristics.

|  | *Support e-cigarettes to transition off cigarettes* |  |  | *prefer patient to use FDA-approved pharmacotherapy rather than e-cigarettes* |  |  | *e-cigarettes are not effective for cessation and discourage their use* |  |  | *e-cigarettes are harmful and discourage their use* |  |  |
| --- | --- | --- | --- | --- | --- | --- | --- | --- | --- | --- | --- | --- |
|  | 29 YO Female, Light Smoker | 65 YO Male, Heavy Smoker | p value | 29 YO Female, Light Smoker | 65 YO Male, Heavy Smoker | p value | 29 YO Female, Light Smoker | 65 YO Male, Heavy Smoker | p value | 29 YO Female, Light Smoker | 65 YO Male, Heavy Smoker | p value |
| Overall | 15.2% | 49.3% | <.0001 | 82.1% | 56.8% | <.0001 | 19.0% | 17.5% | *ns* | 31.7% | 24.9% | <.0001 |
| Cardiology | 23.1% | 53.1% | <.0001 | 71.4% | 50.3% | <.0001 | 15.6% | 15.0% | *ns* | 25.9% | 16.3% | 0.0015 |
| Family Medicine | 15.7% | 49.8% | <.0001 | 85.0% | 61.3% | <.0001 | 19.7% | 18.8% | *ns* | 34.1% | 28.6% | 0.0001 |
| Internal Medicine | 14.3% | 48.8% | <.0001 | 82.4% | 56.3% | <.0001 | 20.0% | 18.7% | *ns* | 32.3% | 23.1% | <.0001 |
| Ob/Gyn | 13.8% | 47.5% | <.0001 | 83.7% | 55.0% | <.0001 | 17.6% | 15.8% | *ns* | 30.9% | 25.8% | 0.0013 |
| Oncology | 12.9% | 46.4% | <.0001 | 83.6% | 60.0% | <.0001 | 16.4% | 17.9% | *ns* | 24.3% | 20.7% | *Ns* |
| Pulmonary | 14.9% | 56.6% | <.0001 | 79.4% | 55.4% | <.0001 | 24.0% | 18.3% | 0.025 | 36.6% | 25.7% | 0.0021 |

\* A 29 year old female cigarette smoker, who smokes 10 cigarettes per day, and has never tried any cessation medications or behavioral counseling for tobacco use
65 year old male cigarette smoker, who smokes 40 cigarettes per day, and has had multiple unsuccessful quit attempts with various FDA-approved cessation medications

## Discussion

Overall, nearly 70% of physician respondents reported being asked about e-cigarettes by their patients and 22% recommended e-cigarettes at one time. These estimates are likely to increase if the popularity of e-cigarettes continues to rise and more importantly, if the evidence that e-cigarettes are effective as cigarette smoking cessation tools strengthens.^10^ The current data reflect increased patient engagement on this issue as the percentage of physicians who had been asked about e-cigarettes in 2019 was greater than 2018.

This study also examined potential clinical scenarios with two different types of patients. Physicians were more likely to recommend e-cigarettes to an older, heavy smoker with multiple failed attempts, yet more likely to advise FDA-approved medications to a younger, lighter smoker who has never tried pharmacotherapy. In other words, when the risks of continued smoking outweighed the potential risk of using e-cigarettes to quit smoking, physicians were more likely to offer an e-cigarette as an option once other options were unsuccessful. In the absence of definitive evidence about e-cigarettes as a cessation tool, it was not surprising that physicians, who often find themselves in situations that may not be directly addressed in standard practice and treatment guidelines, took this pragmatic approach. This finding was consistent with previous qualitative studies showing that physicians were more inclined to recommend e-cigarettes for cessation to certain patients,^24,34^ including those who had multiple unsuccessful quit attempts.^24^ These types of clinical scenarios can simulate what might occur in a true clinical encounter and guide future physician education.

The current findings demonstrated a misperception of the relative harm of e-cigarettes in comparison to other tobacco products, such as combusted tobacco, with 60% of physician respondents believing that all tobacco products were equally harmful. This belief is refuted by the growing body of evidence that combusted tobacco products (e.g., cigarettes and cigars) contribute overwhelmingly to the societal harm of tobacco. While there is no evidence to date suggesting that e-cigarettes are completely harmless, numerous studies suggest that the level of toxins found in these products are orders of magnitude lower than found in combusted tobacco.^2,22,35^ Other studies have demonstrated misperceptions by physicians regarding other important tobacco control issues, such the harm of nicotine.^36,37^ Based on the growing importance of nicotine as a central component of a potential FDA regulatory framework, it is critical to address physicians’ nicotine misperceptions.. In the same respect, harm reduction has become a centerpiece of FDA’s new strategy for a comprehensive nicotine policy which will assess sources of nicotine, based on their relative harm.^38^ It is very important to correct misperceptions regarding the relative harm of various tobacco products as more modified-risk tobacco products (MRTP’s) may be introduced through an FDA authorization process. These results suggest that physician education regarding harm reduction is greatly needed and should be a crucial component of this future strategy. The results also indicated that physicians who already endorse a risk continuum were more likely to recommend e-cigarettes to their patients.

Primary care physicians, pulmonologists, and cardiologists were more likely to be asked about e-cigarettes. Patients may feel reluctant to disclose smoking behavior to their oncology provider due to stigma and guilt.^39^ Physicians who have implemented the PHS guidelines were more likely to be asked about e-cigarettes. This may be a result of patients feeling more comfortable in discussing the issue once it has been raised. Increasing age of the physician was also related to lower rates of being asked about e-cigarettes, possibly due to their perceived lower familiarity with the products in the eye of their patients. Physicians who were ever smokers were more likely to recommend e-cigarettes as perhaps, they best understand the difficulty in quitting.

Physicians who have been asked about e-cigarettes by their patients in the past 30 days were considered “prompted” by their patients. This prompting was by far the strongest predictor for recommending e-cigarettes. This relationship may be a result of physicians’ willingness, in the absence of definitive scientific evidence, to try e-cigarettes as an alternative if the patient has already brought it up. In a prior study, a national sample of physicians were also more likely to engage in evidence-based tobacco treatment if the patient had prompted the treatment from the physician.^40^

Although this study had many strengths - large sample size, diverse physician characteristics, established instruments - there were also some limitations. First, data are self-reported and may be subject to self-report bias. Second, data reduction techniques were used for the logistic regression. Although data reduction increases interpretability, there is the possibility of the loss of granular detail. Third, because sampling probabilities varied by specialty, the overall sample was not nationally representative but was randomly drawn, and analyses were adjusted by specialty.

## Conclusion

With a high number of patients inquiring about e-cigarettes, it is important to understand the perceptions and recommendation practices of physicians as they are a trusted source of health information and are facilitators to tobacco cessation. E-cigarettes will play a pivotal role in FDA’s new nicotine policy framework, and thus will impact tobacco use patterns throughout the country. These findings suggest that some physicians believe e-cigarettes could help patients quit smoking in certain circumstances, but require more evidence regarding their safety and effectiveness. Additionally, physicians’ understanding of e-cigarettes in the context of harm reduction needs to keep pace with the emerging scientific evidence through effective educational opportunities.

## Data Availability

All data produced in the present study are available upon reasonable request to the authors

## Notes

### Competing Interest Statement

The authors have declared no competing interest.

### Funding Statement

This study was funded by the NIH/National Cancer Institute via R01CA190444

### Author Declarations

The Rutgers RBHS Health Sciences IRB committee gave ethical approval for this work and approved the study as exempt.

## REFERENCES

1. Jamal A, Phillips E, Gentzke AS, et al. Current cigarette smoking among adults—United states, 2016. Morb Mortal Weekly Rep. 2018;67(2):53.

2. National Academies of Sciences, Engineering, and Medicine. Public health consequences of e-cigarettes. National Academies Press; 2018.

3. Gentzke AS, Wang TW, Jamal A, et al. Tobacco product use among middle and high school students—United states, 2020. Morb Mortal Weekly Rep. 2020;69(50):1881.

4. Giovenco DP, Lewis MJ, Delnevo CD. Factors associated with e-cigarette use: A national population survey of current and former smokers. Am J Prev Med. 2014;47(4):476–480.

5. Soule EK, Rosas SR, Nasim A. Reasons for electronic cigarette use beyond cigarette smoking cessation: A concept mapping approach. Addict Behav. 2016;56:41–50.

6. Wackowski OA, Bover Manderski MT, Delnevo CD, Giovenco DP, Lewis MJ. Smokers’ early e-cigarette experiences, reasons for use, and use intentions. Tobacco regulatory science. 2016;2(2):133–145.

7. Patel D, Davis KC, Cox S, et al. Reasons for current E-cigarette use among US adults. Prev Med. 2016;93:14–20.

8. Yong H, Borland R, Cummings KM, et al. Reasons for regular vaping and for its discontinuation among smokers and recent ex□smokers: Findings from the 2016 ITC four country smoking and vaping survey. Addiction. 2019;114:35–48.

9. Caraballo RS, Shafer PR, Patel D, Davis KC, McAfee TA. Peer reviewed: Quit methods used by US adult cigarette smokers, 2014–2016. Preventing chronic disease. 2017;14.

10. Hartmann-Boyce J, McRobbie H, Lindson N, et al. Electronic cigarettes for smoking cessation. Cochrane database of systematic reviews. 2021(4).

11. Balfour DJ, Benowitz NL, Colby SM, et al. Balancing consideration of the risks and benefits of E-cigarettes. Am J Public Health. 2021(0):e1–e12.

12. Tobacco, The Clinical Practice Guideline Treating. A clinical practice guideline for treating tobacco use and dependence: 2008 update: A US public health services report. Am J Prev Med. 2008;35(2):158–176.

13. Steinberg MB, Giovenco DP, Delnevo CD. Patient–physician communication regarding electronic cigarettes. Preventive medicine reports. 2015;2:96–98.

14. Nickels AS, Warner DO, Jenkins SM, Tilburt J, Hays JT. Beliefs, practices, and self-efficacy of US physicians regarding smoking cessation and electronic cigarettes: A national survey. Nicotine Tobacco Res. 2017;19(2):197–207.

15. Kanchustambham V, Saladi S, Rodrigues J, Fernandes H, Patolia S, Santosh S. The knowledge, concerns and healthcare practices among physicians regarding electronic cigarettes. Journal of community hospital internal medicine perspectives. 2017;7(3):144–150.

16. Wackowski OA, Bover Manderski MT, Delnevo CD. Smokers’ sources of e-cigarette awareness and risk information. Prev Med Rep. 2015;2:906-910. Accessed Oct 24, 2018. doi: 10.1016/j.pmedr.2015.10.006.

17. Erku DA, Gartner CE, Morphett K, Steadman KJ. Beliefs and self-reported practices of health care professionals regarding electronic nicotine delivery systems: A mixed-methods systematic review and synthesis. Nicotine and Tobacco Research. 2020;22(5):619–629.

18. American cancer society updates position on electronic cigarettes. http://pressroom.cancer.org/eCigs2019. Accessed Aug 31, 2021.

19. AMA calls for total ban on all vaping products not approved by FDA. https://www.ama-assn.org/press-center/press-releases/ama-calls-total-ban-all-vaping-products-not-approved-fda. Accessed Aug 31, 2021.

20. Amos A, Arnott D, Aveyard P, et al. Nicotine without smoke: Tobacco harm reduction.. 2016.

21. McNeill A, Brose LS, Calder R, Bauld L, Robson D. Evidence review of e-cigarettes and heated tobacco products 2018. A report commissioned by public health England.London: Public Health England. 2018;6.

22. Royal College of General Practitioners. RCGP position statement on the use of electronic nicotine vapour products (E-cigarettes). https://www.cancerresearchuk.org/sites/default/files/rcgp_position_statement_on_the_use_of_electronic_nicotine_vapour_products.pdf. Updated 2017.

23. Ofei-Dodoo S, Kellerman R, Nilsen K, Nutting R, Lewis D. Family physicians’ perceptions of electronic cigarettes in tobacco use counseling. The Journal of the American Board of Family Medicine. 2017;30(4):448–459.

24. Singh B, Hrywna M, Wackowski OA, Delnevo CD, Lewis MJ, Steinberg MB. Knowledge, recommendation, and beliefs of e-cigarettes among physicians involved in tobacco cessation: A qualitative study. Preventive medicine reports. 2017;8:25–29.

25. Kandra KL, Ranney LM, Lee JG, Goldstein AO. Physicians’ attitudes and use of e-cigarettes as cessation devices, north carolina, 2013. PloS one. 2014;9(7):e103462.

26. Geletko KW, Myers K, Brownstein N, et al. Medical residents’ and practicing physicians’e-cigarette knowledge and patient screening activities: Do they differ? Health services research and managerial epidemiology. 2016;3:2333392816678493.

27. Egnot E, Jordan K, Elliott JO. Associations with resident physicians’ early adoption of electronic cigarettes for smoking cessation. Postgrad Med J. 2017;93(1100):319–325.

28. Baldassarri SR, Chupp GL, Leone FT, Warren GW, Toll BA. Practise patterns and perceptions of chest health care providers on electronic cigarette use: An in-depth discussion and report of survey results. Journal of smoking cessation. 2018;13(2):72–77.

29. England LJ, Anderson BL, Mahoney J, Coleman-Cowger VH, Melstrom P, Schulkin J. Screening practices and attitudes of obstetricians-gynecologists toward new and emerging tobacco products. Obstet Gynecol. 2014;211(6):695. e1–695. e7.

30. Northrup TF, Klawans MR, Villarreal YR, et al. Family physicians’ perceived prevalence, safety, and screening for cigarettes, marijuana, and electronic-nicotine delivery systems (ENDS) use during pregnancy. The Journal of the American Board of Family Medicine. 2017;30(6):743–757.

31. Delnevo CD, Singh B. The effect of a web-push survey on physician survey responses rates: A randomized experiment. Survey practice. 2021;14(1).

32. American Association for Public Opinion Research. Standard definitions: Final dispositions of case codes and outcome rates for surveys. 2011.

33. Steinberg MB, Delnevo CD. Physician beliefs regarding effectiveness of tobacco dependence treatments: Results from the NJ health care provider tobacco survey. Journal of general internal medicine. 2007;22(10):1459–1462.

34. El-Shahawy O, Brown R, Elston Lafata J. Primary care physicians’ beliefs and practices regarding e-cigarette use by patients who smoke: A qualitative assessment. International journal of environmental research and public health. 2016;13(5):445.

35. Goniewicz ML, Gawron M, Smith DM, Peng M, Jacob P, Benowitz NL. Exposure to nicotine and selected toxicants in cigarette smokers who switched to electronic cigarettes: A longitudinal within-subjects observational study. Nicotine Tobacco Res. 2017;19(2):160–167.

36. Steinberg MB, Manderski MTB, Wackowski OA, Singh B, Strasser AA, Delnevo CD. Nicotine risk misperception among US physicians. Journal of General Internal Medicine. 2020:1–3.

37. Bover Manderski MT, Steinberg MB, Wackowski OA, Singh B, Young WJ, Delnevo CD. Persistent misperceptions about nicotine among US physicians: Results from a randomized survey experiment. International journal of environmental research and public health. 2021;18(14):7713.

38. Gottlieb S, Zeller M. A nicotine-focused framework for public health. N Engl J Med. 2017;377(12):1111–1114.

39. Simmons VN, Litvin EB, Patel RD, et al. Patient–provider communication and perspectives on smoking cessation and relapse in the oncology setting. Patient Educ Couns. 2009;77(3):398–403.

40. Steinberg MB, Akincigil A, Delnevo CD, Crystal S, Carson JL. Gender and age disparities for smoking-cessation treatment. Am J Prev Med. 2006;30(5):405–412.

